# A novel approach for LV delay evaluation: non-invasive QLV measurement using UHF-ECG

**DOI:** 10.1101/2025.09.17.25336029

**Authors:** Lukáš Povišer, Petr Štros, Óscar Cano, Pavel Leinveber, Radovan Smíšek, Jana Veselá, Karol Čurila

## Abstract

**Introduction:** Ultra-high-frequency ECG (UHF-ECG) which displays ventricular dyssynchrony may be a valuable tool for improving the selection and treatment of patients with indication to cardiac resynchronization therapy (CRT).

**Objective:** To compare noninvasive assessment of LV delay using UHF-ECG with invasive QLV in CRT, and to evaluate the effect of biventricular CRT on LV resynchronization and remodeling.

**Methods:** Consecutive patients undergoing CRT at two centers were included. LV lead positions were classified as basal, midventricular, or apical lateral wall. QLV was measured from QRS onset to the steepest EGM deflection. UHF-ECG LV delay was measured from QRS onset to local activation in leads V5-V8. LV dyssynchrony was calculated as the time from the earliest activation to the latest activation in leads V4–V8.

**Results:** 48 patients were included (75 % male, age 67 ± 12 years, LVEF 31 ± 6 %, QRSd 167 ± 20 ms). UHF-ECG activation delay in V8 did not differ from QLV in the basal segment (3 ± 14 ms, *p* = 0.52). QLV from the midventricular segment did not differ from UHF-ECG activation in V7 (−3 ± 17 ms, *p* = 0.18). QLV from the apical segment was not different from UHF-ECG activation in V6 (0 ± 17 ms, *p* = 0.92).

Patients with QLV and lv-DYS above 95 and 60 ms showed a more significant reduction in LV dyssynchrony and were more often responders to CRT than patients with shorter values (Δlv-DYS -70 ms vs. -8 ms, and responders 95 % vs. 24 %).

**Conclusion:** UHF-ECG enables noninvasive estimation of QLV and assessment of CRT response, which may have important clinical implications for the evaluation and management of CRT candidates.

**Graphical abstract:** 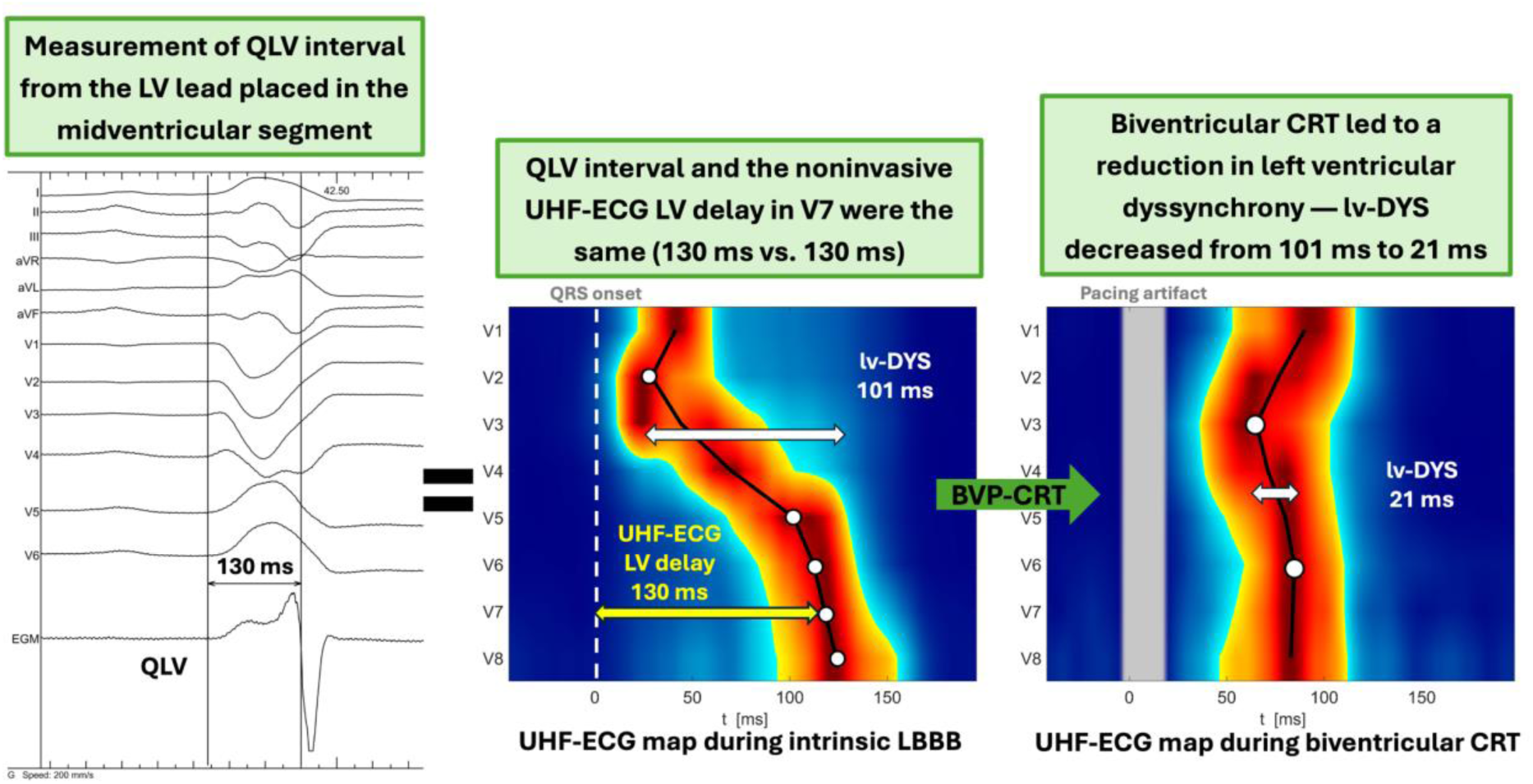

## Introduction

Cardiac resynchronization therapy (CRT) has been demonstrated to improve quality of life, enhance left ventricular (LV) systolic function, and reduce heart failure (HF) hospitalizations and mortality in patients with ventricular electrical dyssynchrony.^1–3^ Recent studies have shown that LV pacing at sites with prolonged electrical delay (LV delay) offers the greatest potential to correct ventricular dyssynchrony and improve clinical response in CRT patients with non-RBBB morphology.^4–7^

The degree of LV delay is routinely assessed using the QLV interval, defined as the time from the onset of the QRS complex on the surface ECG to the steepest deflection of the local electrogram recorded from the implanted LV lead positioned in one of the coronary sinus branches. Notably, a QLV value greater than 95 ms has been identified as one of the most powerful predictors of a favorable response to CRT.^6,8–10^

Over the last few years, a novel method of non-invasive ventricular dyssynchrony assessment using ultra-high-frequency (UHF-ECG) has been studied.^11–14^ UHF-ECG enables visualization of the ventricular activation sequence and allows precise quantification of dyssynchrony by measuring time delays between activations of different LV segments using the standard chest leads (V1–V8). However, the relationship between LV electrical delay measured invasively as QLV and noninvasively using UHF-ECG remains unknown.

This study aimed to compare QLV with noninvasively assessed LV delay obtained by UHF-ECG, and to evaluate the changes in LV synchrony and LV remodeling after biventricular pacing in CRT candidates.

## Methods

This retrospective study included patients with symptomatic heart failure and an indication for CRT. The inclusion criteria were (1) symptomatic heart failure with LVEF ≤ 40 %, (2) spontaneous rhythm with a non-RBBB morphology and QRS duration (QRSd) ≥ 130 ms, and (3) successful coronary sinus cannulation with measurements of QLV in one of its branches.

### Implantation procedure and the LV lead positioning

CRT device implantation was performed at two participating centers. All patients provided written informed consent prior to the procedure, and the study was conducted in accordance with the principles outlined in the Declaration of Helsinki.

Following venous access via the left subclavian vein, the right ventricular lead was typically placed in the apico-septal region. The coronary sinus (CS) was then cannulated, and a quadripolar or bipolar LV lead (Boston Scientific, USA) was advanced into a suitable lateral, posterolateral, or anterolateral CS branch. In cases with challenging venous anatomy, a guidewire alone (Boston Scientific, USA) was used to acquire local LV electrograms (EGMs) without permanent lead implantation.

After successful lead positioning, each lead electrode was connected to a recording system, and local LV EGMs were acquired. Simultaneously, biplane fluoroscopic images (left anterior oblique (LAO) 30°, right anterior oblique (RAO) 30°) were obtained to document the anatomical position of the LV lead. In most patients, the LBBAP lead was simultaneously implanted into the septum as a part of the research project comparing LV hemodynamics between BVP and LBBAP,^15^ and at the end of the procedure the LV lead was extracted and LBBAP was utilized as a CRT method.

### Invasive QLV measurements

The unipolar left ventricular electrogram (LV EGM) recorded from the implanted LV lead and the surface 12-lead ECG were displayed simultaneously at a sweep speed of 200 mm/s on an electrophysiological recording system, Labsystem Pro (Boston Scientific, USA). The QLV interval was retrospectively measured using electronic calipers from the onset of the native QRS complex on the surface 12-lead ECG to the local activation on LV EGM, as illustrated in Figure 1. The local activation time was defined as the point of the steepest negative deflection on the unipolar LV EGM, corresponding to the maximum negative derivative (dV/dt_min_). For each LV lead electrode, the QLV interval was measured on three consecutive beats, and the average of these measurements was used as the final value. In every patient, the longest measured QLV interval was identified.

**Figure 1:**
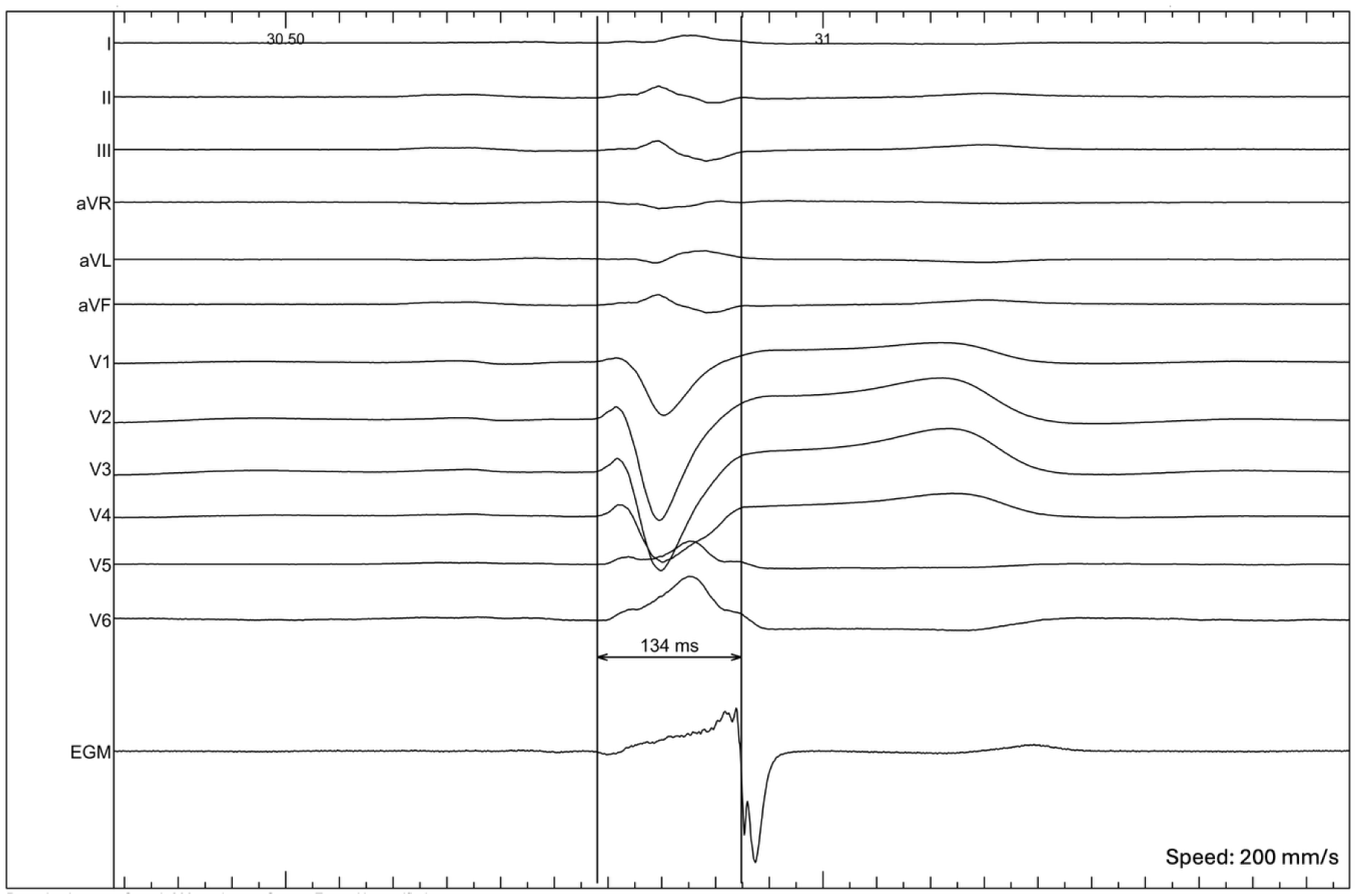
Simultaneous display of a surface 12-lead ECG and a unipolar LV EGM. The QLV interval is manually measured using electronic calipers from the onset of the QRS complex on the surface ECG to the steepest negative deflection on the LV EGM.

### Assessment of LV lead placement across ventricular segments

During CRT implantation, the final LV lead position was documented using fluoroscopic imaging in two standard projections: left anterior oblique (LAO 30°) and right anterior oblique (RAO 30°). Based on these projections, the lateral wall of the left ventricle was divided longitudinally into basal, midventricular, and apical segments, and circumferentially into anterior, lateral, and inferior segments, as illustrated in Figure 2. Subsequently, each electrode of the LV lead was then retrospectively assigned to one of these anatomical regions. This segment classification was used for further analysis of QLV intervals and UHF-ECG activation delay in relation to lead location.

**Figure 2:**
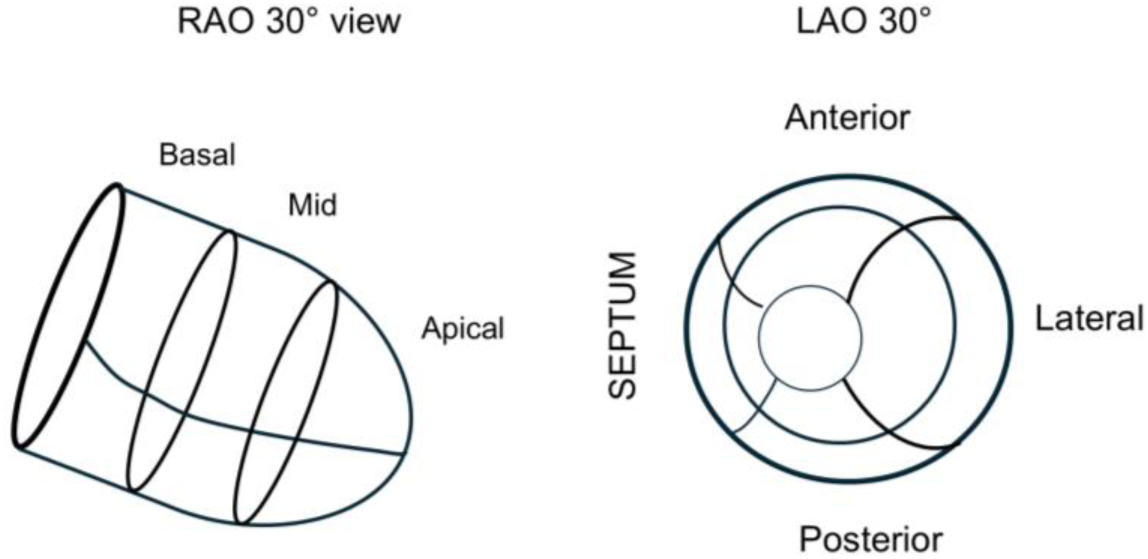
Basal, midventricular and apical segments of the left ventricle are shown in the right anterior oblique 30° view. Anterior, lateral and posterior segments of the left ventricle are shown in the left anterior oblique 30° view.

### UHF-ECG data acquisition and analysis

In all patients, two 5-minute 14-lead UHF-ECG recordings were obtained using an ultra-high-frequency ECG acquisition system (ISI, Brno, Cardion, FNUSA, CZ, 2018). Signal processing and UHF-ECG map generation followed the previously published methodology and were consistent with earlier studies.^11–14^ In brief, median amplitude envelopes were computed for 16 frequency bands (150–1000 Hz) for each chest lead. The broadband QRS complex (UHF-QRS) was constructed as the average of the 16 normalized median amplitude envelopes and displayed as a color map for each lead. Local myocardial activation times were calculated as the center of mass of UHF-QRS above 50% threshold of the baseline-to-peak amplitude in each chest lead. UHF-ECG recordings during spontaneous rhythm were performed before the CRT procedure, while biventricular paced rhythm was recorded after implantation at a fixed heart rate of 100 bpm and a VV delay of 0 ms. Standard V1–V8 chest lead positions were used.

To quantify UHF-ECG LV activation delay during the spontaneous rhythms, the time from the QRS onset to the local activation centers in leads V5, V6, V7, and V8 was measured (Figure 3A). To quantify LV dyssynchrony both during the spontaneous and paced rhythms, the index of LV electrical dyssynchrony (lv-DYS) was calculated as the difference between the earliest UHF-ECG local myocardial activation among leads V1–V8 and the latest local myocardial activation in leads V4–V8 – Figure 3B.

**Figure 3:**
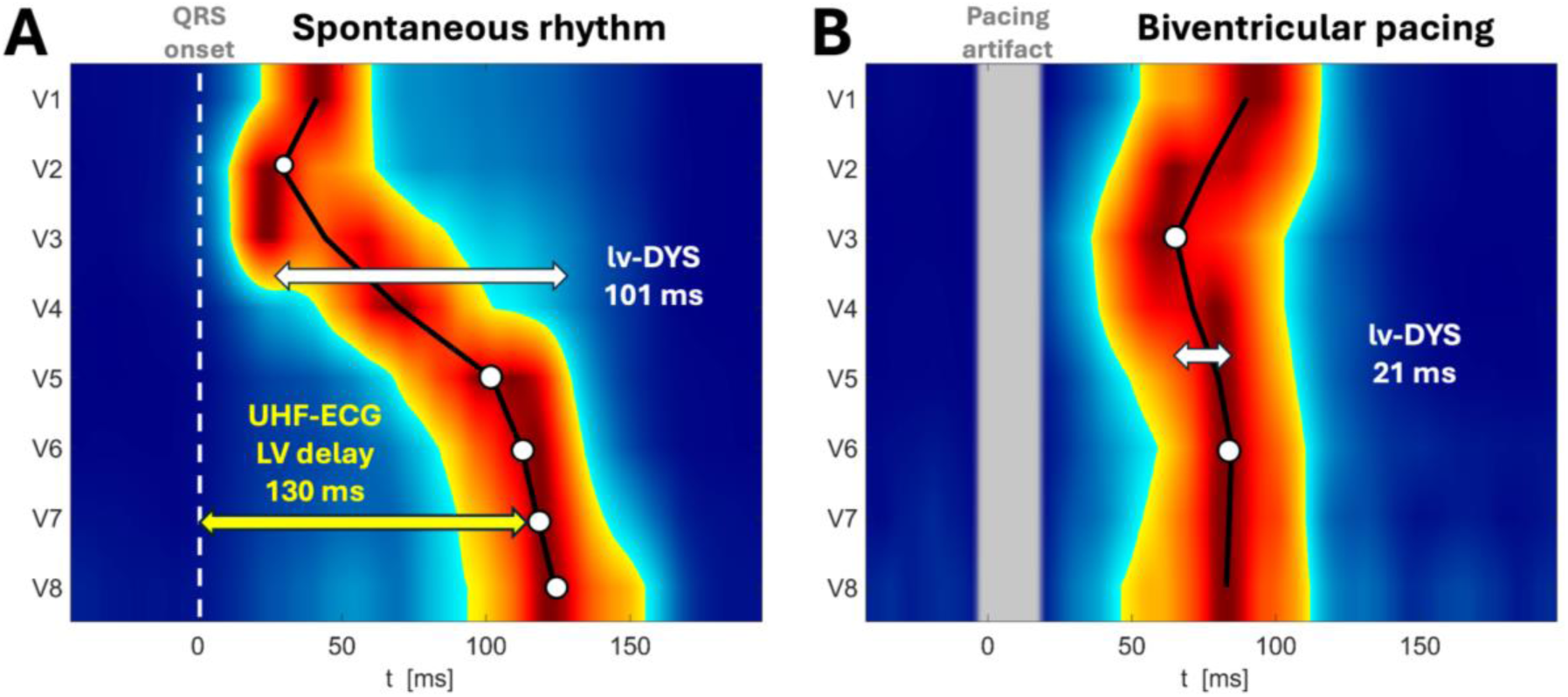
**Panel A** – UHF-ECG map during intrinsic conduction with measurement of UHF-ECG LV delay in lead V7 (time interval from QRS onset (placed on 0 ms) to local myocardial activation under the lead). UHF-ECG LV delay can only be measured during spontaneous rhythms, as it is referenced to the onset of the intrinsic QRS complex. **Panel B** – UHF-ECG map during biventricular pacing (BVP-CRT), showing measurement of lv-DYS (the interval from the first local UHF-ECG activation to the latest activation among leads V4–V8). lv-DYS can be assessed during both spontaneous and paced rhythms, as it is measured from the earliest to the latest UHF-ECG activation and is independent of QRS onset.

#### Echocardiographic outcomes after BVP-CRT

To understand if UHF-ECG left ventricular dyssynchrony during spontaneous rhythm plays a role in left ventricular remodeling after BVP-CRT, we studied a historical cohort of 42 patients with heart failure. All of them had UHF-ECG recording of the spontaneous rhythm before an implant procedure, LVEF ≤ 35% and QRS complex of non-RBBB morphology lasting ≥ 130 ms. Echocardiography was done before and on average 6 months after BVP-CRT. CRT response was defined as a reduction in left ventricular end-systolic volume of at least 10 %.

#### Statistical analysis

Mean activation delay, mean differences, and standard deviations were calculated to identify the UHF-ECG lead with the closest agreement to QLV within each anatomical LV segment. The strength of associations between QLV and the best-matching UHF-ECG leads was evaluated using Pearson’s correlation coefficient (r). Scatter plots with fitted linear regression lines were used for visualization.

The Kolmogorov–Smirnov test was used to assess normality. Depending on the distribution, comparisons between paired values were performed using either the paired t-test (for normally distributed data) or the Wilcoxon signed-rank test (for non-normally distributed data). Categorical data are presented as absolute numbers and percentages. All statistical analyses were performed using MATLAB (MATLAB2023b, The MathWorks Inc, Natick, MA, USA).

## Results

### Characteristics of study subjects

A total of 48 patients (75 % males) with a mean age of 67 ± 12 years were included in the study. The average left ventricular ejection fraction (LVEF) was 31 ± 6 %, and the mean left ventricular end-diastolic diameter (LVEDD) was 61 ± 7 mm. Ischemic cardiomyopathy was present in 18 patients (38 %). Six patients (13 %) presented with non-specific intraventricular conduction delay (NIVCD), while 42 patients (87 %) fulfilled the Strauss criteria for left bundle branch block (LBBB). The average intrinsic QRSd was 167 ± 20 ms. Additional clinical characteristics are summarized in Table 1.

**Table 1:** Clinical characteristics of patients.

|  |  |
| --- | --- |
| Age (years), mean ± SD | $67 \pm 12$ |
| Male gender, n (%) | 36 (75) |
| Comorbidities: |  |
| • Coronary artery disease, n (%) | 18 (38) |
| • Diabetes mellitus, n (%) | 17 (35) |
| • Hypertension, n (%) | 33 (69) |
| LV ejection fraction (%), mean ± SD | $31 \pm 6$ |
| LVEDD (mm), mean ± SD | $61 \pm 7$ |
| QRS morphology |  |
| • LBBB, n (%) | 42 (87) |
| • NIVCD, n (%) | 6 (13) |
| Intrinsic QRSd (ms), mean ± SD | $167 \pm 20$ |

### Comparison of QLV and UHF-ECG activation delay across LV segments

Of the total number of 174 LV lead electrodes, 105 (60 %) were located in the basal segment of the LV, 54 (31 %) in the mid-segment and 15 (9 %) in the apical segment. All analyzed LV leads were placed in the lateral branches of the CS, none were in the anterior or posterior branches. As shown in the **Figure 4A**, the average QLV was longest in the basal LV segment and shortest in the apical segment (122 ± 24 ms vs. 107 ± 21 ms, *p* = 0.04). The average UHF-ECG LV delay was longest in the lead V8 and shortest in the lead V5 (121 ± 25 ms vs. 102 ± 21 ms, *p* < 0.01) – **Figure 4B**.

**Figure 4:**
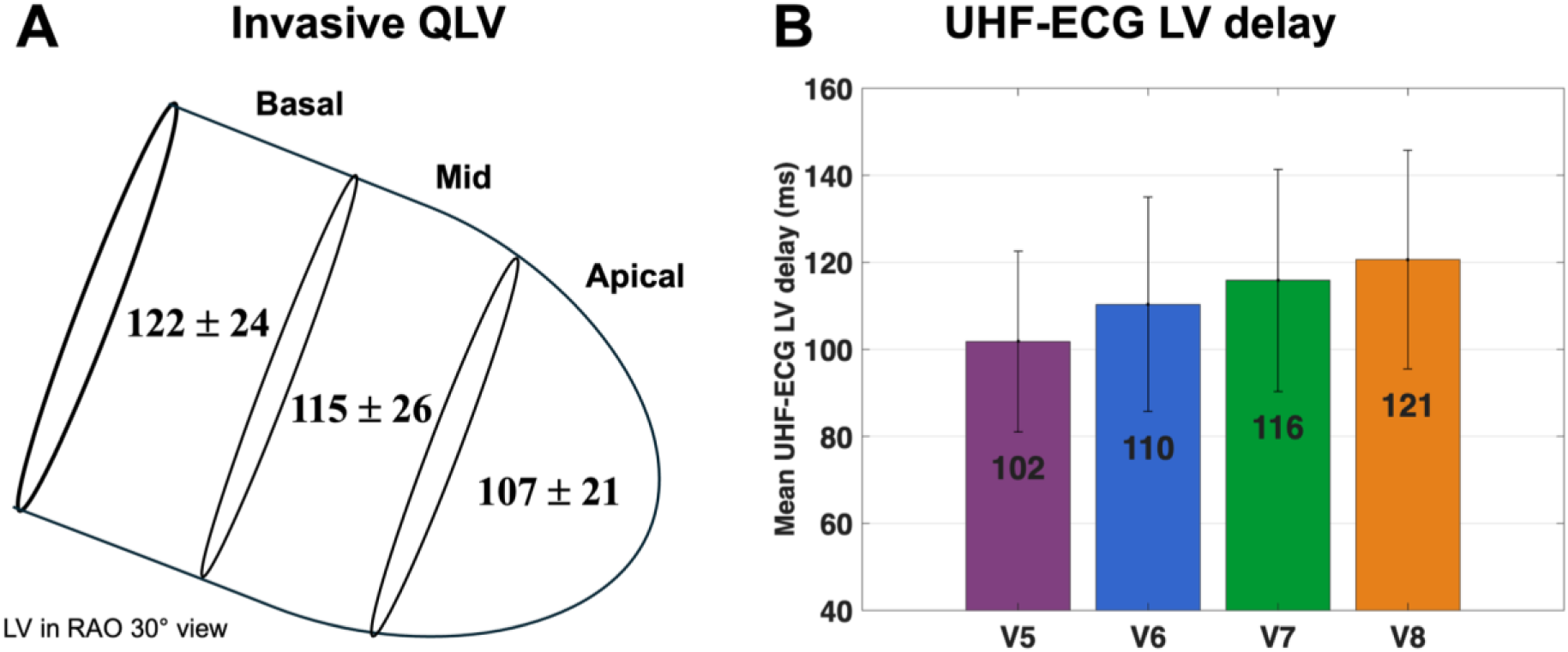
**Panel A** - Mean QLV from the basal, mid and apical segments of the LV. **Panel B** - UHF ECG activation delay in leads V5, V6, V7, and V8.

No difference was observed between UHF-ECG activation delay in lead V8 and QLV measured in the **basal segment** (mean difference 3 ± 14 ms, *p* = 0.52). For the **midventricular segment**, no difference was found between UHF-ECG activation delays in leads V7 and V8 and QLV (mean difference: –3 ± 17 ms for V7, *p* = 0.18 and 4 ± 14 ms for V8, *p* = 0.19). For the **apical segment**, UHF-ECG activation delays in leads V6 and V7 showed no difference compared to QLV (mean difference: 0 ± 17 ms for V6, *p* = 0.92; 6 ± 16 ms for V7, *p* = 0.16) – **Figure 5A**. In one patient which underwent a chest MRI scan with attached ECG leads, we were able to demonstrate that V6-V8 chest leads projections onto the closest epicardial region covered the inferolateral basal-to-apical LV region – **Figure 5B**.

**Figure 5:**
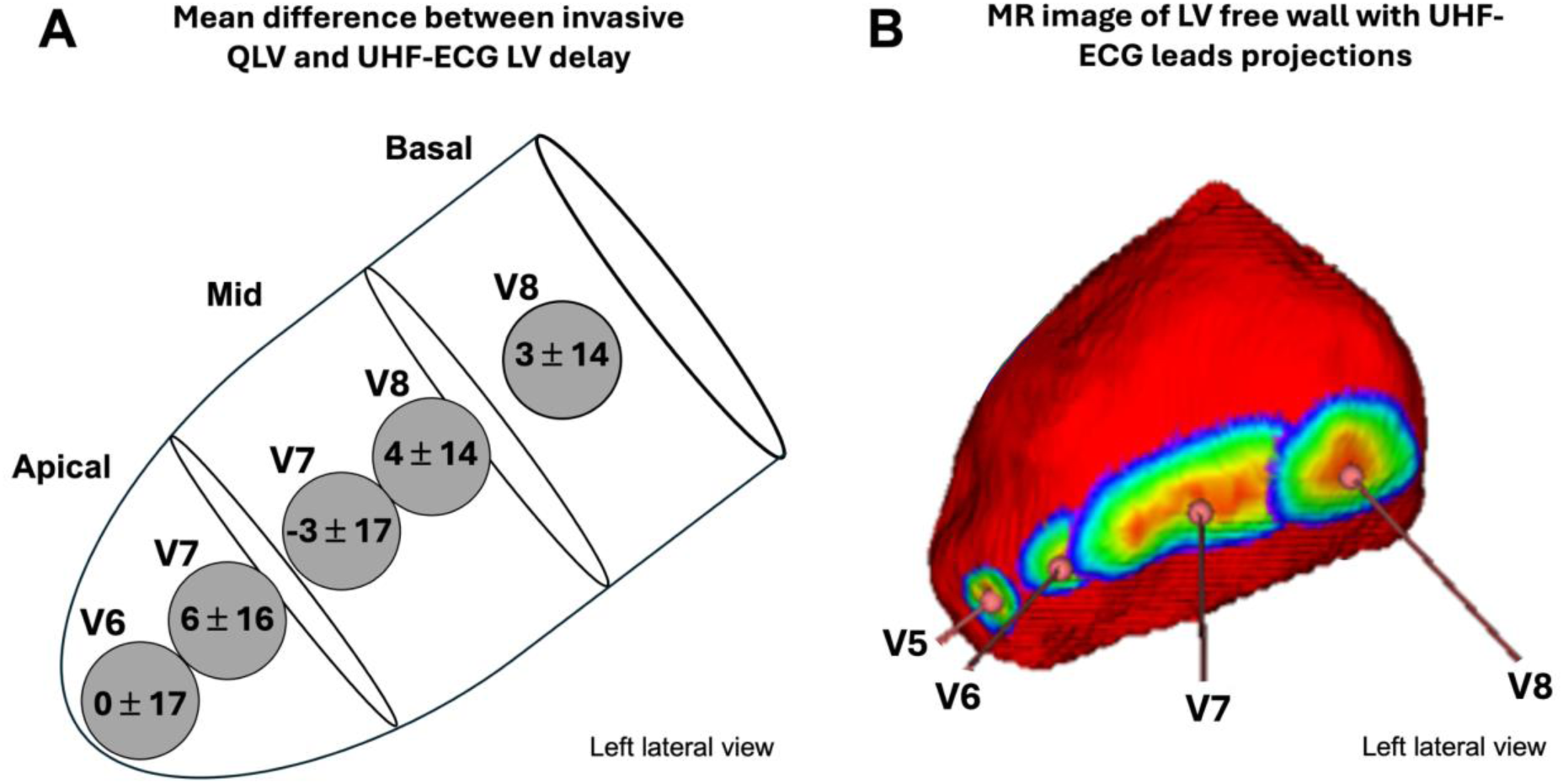
**Panel A** - Left lateral view of the left ventricle showing the mean difference between invasive QLV for each LV segment and UHF-ECG LV activation delay for leads V6-V8. Only leads with a statistically non-significant difference compared to QLV are displayed. **Panel B -** Magnetic resonance image of the LV free wall in left lateral view showing the projection of UHF-ECG leads V5–V8 onto the anatomically closest regions of the LV epicardium in a patient with an indication for CRT. The shortest distance between the lead and the LV epicardium was defined as the center of gravity of the region where the variation in distance was less than 5 %. Colors within this region represent the relative distance from the lead (red indicating the shortest, and blue the longest distance).

UHF-ECG activation delays in other leads significantly differed from QLV, mean differences are shown in the **Table 2**. A strong linear correlation between QLVs in any LV segment and UHF-ECG LV delays for leads V6-V8 (r = 0.84) was observed – **Figure 6**.

**Figure 6:**
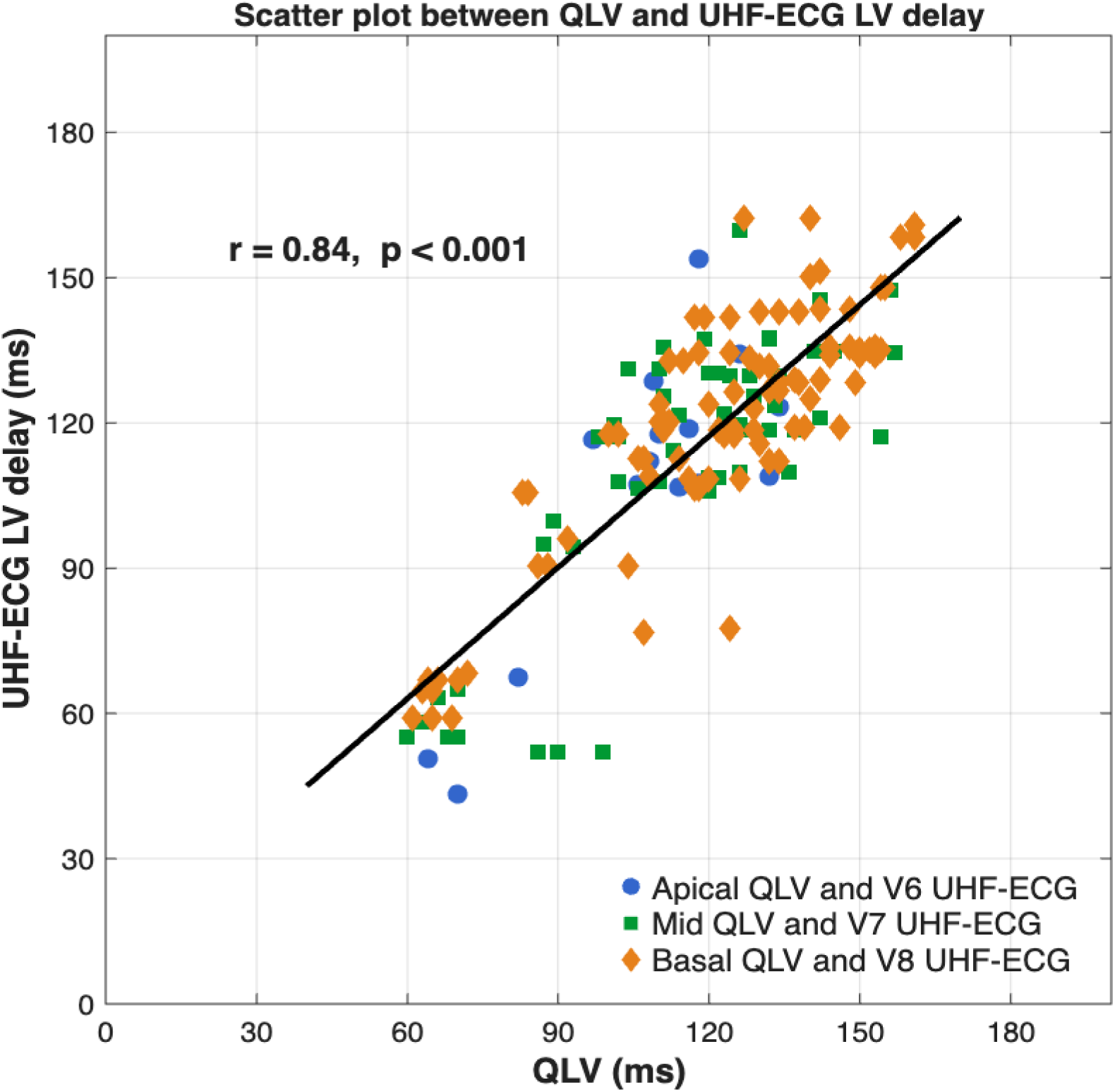
Scatter plot with linear regression between QLV and UHF-ECG LV delay (apical QLV vs. V6 lead, mid QLV vs. V7 lead and basal QLV vs. V8 lead)

**Table 2:** Mean difference between QLV from basal, mid and apical segments and UHF-ECG LV delay in V5, V6, V7 and V8 leads.

|  | V5 | V6 | V7 | V8 |
| --- | --- | --- | --- | --- |
| **Basal QLV** | - 21 ± 17<br>(*p* < 0.001) | - 12 ± 16<br>(*p* < 0.001) | - 6 ± 14<br>(*p* < 0.001) | **3 ± 14**<br>(*p* = 0.52) |
| **Mid QLV** | -16 ± 16<br>(*p* < 0.001) | - 9 ± 17<br>(*p* < 0.001) | **-3 ± 17**<br>(*p* = 0.18) | **4 ± 14**<br>(*p* = 0.19) |
| **Apical QLV** | -8 ± 15<br>(*p* < 0.001) | **0 ± 17**<br>(*p* = 0.92) | **6 ± 16**<br>(*p* = 0.16) | 12 ± 13<br>(*p* < 0.001) |

### Assessment of electrical resynchronization after biventricular CRT using UHF-ECG

To evaluate LV dyssynchrony during both spontaneous and paced rhythms, we assessed lv-DYS as the earliest local activation in leads V1–V8 and the latest activation in leads V4-V8 – **Figure 3**.

During intrinsic conduction, the mean lv-DYS was 82 ± 29 ms, and the mean longest QLV was 125 ± 26 ms. A strong linear correlation was observed between the lv-DYS and the longest QLV (r = 0.76) – **Figure 7A**.

**Figure 7:**
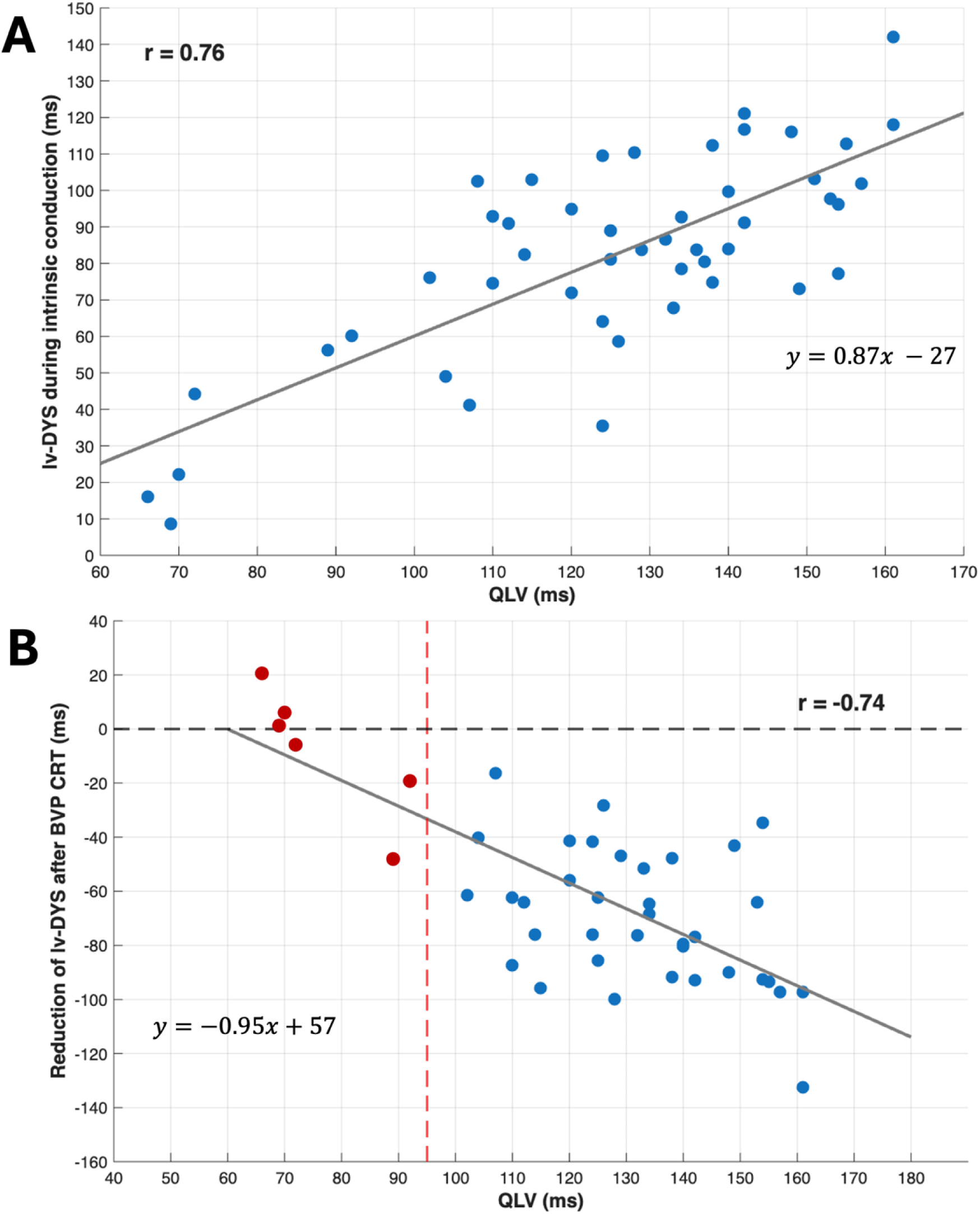
**Panel A** - Scatter plot with linear regression of QLV and lv-DYS during intrinsic conduction. **Panel B -** Scatter plot with linear regression showing the association between the longest QLV for each patient and the reduction in lv-DYS following BVP-CRT. The vertical dashed line at 95 ms separates patients into two groups – in red are patients with the longest QLV below 95 ms, and in blue are patients with the longest QLV above 95 ms.

BVP-CRT resulted in a reduction of lv-DYS compared to intrinsic conduction (21 ± 12 ms vs. 82 ± 29 ms, *p* <0.001). In patients with the longest QLV below 95 ms, the mean lv-DYS did not change after BVP-CRT compared to intrinsic conduction (35 ± 22 ms vs. 27 ± 15 ms, *p* = 0.84). In contrast, in patients with QLV above 95 ms, the mean lv-DYS after BVP-CRT was significantly reduced compared to intrinsic conduction (19 ± 11 ms vs. 89 ± 21 ms, *p* < 0.001) – **Figure 7B**.

An lv-DYS value of 60 ms during spontaneous rhythm was found to separate patients with QLV below 95 ms and above 95 ms – **Figure 7A**.

### Baseline lv-DYS for prediction of CRT response

In a historical cohort of 42 CRT patients, the mean baseline lv-DYS during spontaneous rhythm was 68 ± 29 ms. Patients with lv-DYS above 60 ms showed a greater reduction in ESV at 6 months after CRT compared to those with lv-DYS below 60 ms (24 ± 13 % vs. 4 ± 11 %, *p* < 0.001) and were more often responders (95 % vs. 24 %).

## Discussion

In our study, we found that the invasively acquired parameter QLV can be reliably assessed noninvasively using UHF-ECG. Furthermore, we found that in patients with a shorter QLV and lv-DYS (below 95 ms and 60 ms, respectively), biventricular CRT did not reduce left ventricular dyssynchrony. We also observed that in patients with QLV and lv-DYS above 95 ms and 60 ms, biventricular CRT not only led to a significant reduction in left ventricular dyssynchrony, but also to greater LV remodeling, and a much higher response rate to biventricular CRT compared to patients with shorter values.

### Non-invasive measurement of LV delay vs. invasive QLV measurement

Assessing LV delay using pacing leads placed via the coronary venous system and measuring QLV is an established technique for evaluating electrical left ventricular dyssynchrony during intrinsic conduction. Over the past two decades, several studies have demonstrated that patients with longer QLV intervals are more likely to respond to CRT.^4,6–9^ These patients show greater acute hemodynamic improvement, more pronounced LV reverse remodeling, reduced risk of heart failure hospitalization, and improved long-term survival compared to patients with shorter QLV. However, this invasive approach is inherently limited by anatomical variability in the venous structures, procedural complexity, and its availability being restricted to implantation procedures. For these reasons, a non-invasive approach to LV delay assessment would be highly desirable.

To date, the only published study that directly compared a noninvasive method with invasive QLV is by Melgaard et al. (2022). Their method differs from ours in that it uses an inverse ECG algorithm combined with a generic geometric heart model and the standard 12-lead ECG to estimate the activation sequence and QLV at specific locations in LBBB patients.^16^ In contrast, our method directly measures local myocardial activation under the ECG electrodes. Both methods demonstrated similar accuracy: in the basal segment, their error was 5 ± 16 ms compared to our 3 ± 14 ms; in the mid segment, −10 ± 23 ms vs. −3 ± 17 ms, and in the apical segment, −13 ms vs. 0 ± 17 ms. However, the drawback of their method is its complexity, making it suitable primarily for research settings and limiting its applicability in routine clinical practice. By contrast, UHF-ECG is a noninvasive point-of-care tool that is nowadays easily implemented in everyday clinical workflows worldwide.

### LV dyssynchrony reduction and response to BVP-CRT

The primary goal of CRT is to reduce left ventricular dyssynchrony, as this is the key mechanism leading to clinical benefit in responding patients.^17^ Aside from QRS duration, there are currently no widely available clinical tools to assess whether dyssynchrony has been effectively corrected during biventricular pacing. Previous studies have shown that patients with short QLV intervals (below 95 ms) do not respond to CRT as well as patients with longer QLV intervals (above 95 ms).^4,6–9^ Our results corroborate these findings and provide further explanation, i.e., patients with short QLV also present with small LV dyssynchrony at baseline, which does not improve after biventricular CRT and may be a surrogate for nonresponse. In contrast, patients with QLV above 95 ms exhibit pronounced LV dyssynchrony, which significantly decreases during biventricular CRT and leads to a response.

### Clinical application

Noninvasive assessment of electrical LV lateral wall delay and LV dyssynchrony using UHF-ECG provides valuable information before the implantation procedure. This may help the implanting physician better understand which patients are likely to respond or not respond to CRT, optimize patient selection, and predict the clinical course of CRT candidates.

### Limitations

Knowing the exact location and orientation of the heart using 3D imaging, combined with the precise placement of the UHF-ECG electrodes on the chest, would likely improve consistency between QLV and UHF-ECG measurements across patients. However, 3D imaging is not routinely performed in CRT candidates at our institutions. Additionally, some differences in the measurements may result from patient anatomy and variations in how each staff member places UHF-ECG electrodes.

## Conclusions

QLV can be estimated noninvasively using UHF-ECG in patients with non-RBBB morphology undergoing CRT. Non-invasive assessment of LV synchrony by UHF-ECG provides a potential explanation for why patients with shorter QLV values do not respond to biventricular CRT as well as patients with longer QLV values. These findings suggests that non-invasive assessment of dyssynchrony by UHF-ECG may have important clinical implications for the evaluation and management of CRT candidates.

## Data Availability

Available upon request from the authors

## Abbreviations

CRT: cardiac resynchronization therapy
LV: left ventricle
CS: coronary sinus
EGM: electrogram
QLV: QRS onset to Left Ventricular electrogram
BVP: biventricular pacing
UHF-ECG: ultra-high-frequency electrocardiogram
lv-DYS: parameter of LV electrical dyssynchrony
ESV: end systolic volume

